# Analysis and validation of a highly sensitive one-step nested quantitative real-time polymerase chain reaction assay for specific detection of severe acute respiratory syndrome coronavirus 2

**DOI:** 10.1101/2020.08.27.20182832

**Authors:** Yang Zhang, Chunyang Dai, Huiyan Wang, Yong Gao, Tuantuan Li, Yan Fang, Zuojun Shen, Lichang Chen, Zhaowu Chen, Xuejun Ma, Ming Li

**Affiliations:** Department of Clinical Laboratory, The First Affiliated Hospital of University of Science and Technology of China, Hefei 230031, China; Department of Clinical Laboratory, Fuyang Second People’s Hospital, Fuyang Infectious Disease Clinical College, Anhui Medical University, Fuyang 236015, China; National Institute for Viral Disease Control and Prevention, Chinese Center for Disease Control and Prevention, Beijing 102206, China; Center for Biosafety Mega-Science, Chinese Academy of Sciences, Wuhan 430071, China

**Keywords:** COVID-19, SARS-CoV-2, qRT-PCR, OSN-qRT-PCR, ddPCR, highly sensitive

## Abstract

Coronavirus disease 2019 (COVID-19), caused by SARS-CoV-2, is posing a serious threat to global public health. Reverse transcriptase real-time quantitative polymerase chain reaction (qRT-PCR) is widely used as the gold standard for clinical detection of SARS-CoV-2. Due to technical limitations, the reported positive rates of qRT-PCR assay of throat swab samples vary from 30%–60%. Therefore, the evaluation of alternative strategies to overcome the limitations of qRT-PCR is required. A previous study reported that one-step nested (OSN)-qRT-PCR revealed better suitability for detecting SARS-CoV-2. However, information on the analytical performance of OSN-qRT-PCR is insufficient. In this study, we aimed to analyze OSN-qRT-PCR by comparing it with droplet digital PCR (ddPCR) and qRT-PCR by using a dilution series of SARS-CoV-2 pseudoviral RNA and a quality assessment panel. The clinical performance of OSN-qRT-PCR was also validated and compared with ddPCR and qRT-PCR using specimens from COVID-19 patients. The LoD (copies/ml) of qRT-PCR, ddPCR, and OSN-qRT-PCR were 520.1 (95% CI): 363.23–1145.69) for ORF1ab and 528.1 (95% CI: 347.7–1248.7) for N, 401.8 (95% CI: 284.8–938.3) for ORF1ab and 336.8 (95% CI: 244.6–792.5) for N, and 194.74 (95% CI: 139.7–430.9) for ORF1ab and 189.1 (95% CI: 130.9–433.9) for N, respectively. Of the 34 clinical samples from COVID-19 patients, the positive rates of OSN-qRT-PCR, ddPCR, and qRT-PCR were 82.35% (28/34), 67.65% (23/34), and 58.82% (20/34), respectively. In conclusion, the highly sensitive and specific OSN-qRT-PCR assay is superior to ddPCR and qRT-PCR assays, showing great potential as a technique for detection of SARS-CoV-2 in patients with low viral loads.

## INTRODUCTION

The global coronavirus disease 2019 (COVID-19) pandemic, caused by infection of severe acute respiratory syndrome coronavirus 2 (SARS-CoV-2), is posing an enormous burden on social, economic, and healthcare systems worldwide^[1]^. As there is currently no specific treatment option, early detection of SARS-CoV-2-infected patients has facilitated effective isolation and treatment to prevent disease spread. Currently, clinical diagnosis of COVID-19 is mainly confirmed by detecting SARS-CoV-2 RNA using reverse transcriptase real-time quantitative polymerase chain reaction (qRT-PCR)^[2-5]^. However, the sensitivity and reliability of qRT-PCR has been questioned due to cases of negative results in some patients who were highly suspected of having the disease based on clinical presentation and exposure history [6,7]

Droplet digital PCR (ddPCR) is a third generation PCR based on the principles of limited dilution and Poisson statistics^[8,9]^, which works by separating a sample into thousands to millions of droplets and then partitioning them to be read as either positive or negative depending on fluorescence amplitude^[10–13]^. These vast and highly consistent oil droplets substantially improve the detection dynamic range and accuracy of ddPCR^[14]^. In recent years, ddPCR has found many applications, such as analysis of viral load from clinical samples, detection of rare mutations, analysis of copy number variation (CNV), and precise miRNA quantification ^[15-17]^

Given the high sensitivity of ddPCR, Zhao JK et al. utilized this technique to evaluate the viral loads of SARS-CoV-2 from upper respiratory tract specimens for the first time, showing that ddPCR can accurately reflect the viral loads of such specimens, especially nasopharyngeal swabs^[18]^. Subsequently, Renfei Lu et al. used serial dilutions of the same clinical samples to demonstrate that the LoD of ddPCR is at least 10 times better than that of qRT-PCR^[19]^. However, the limitation of the ddPCR assay is that it often needs unique supporting reagents, instruments, and professional operators, causing high running costs with moderate throughput. More convenient and sensitive methods are urgently needed as alternative diagnostic approaches for detecting SARS-CoV-2.

Nested PCR typically utilizes two sequential amplification reactions, each of which uses a different pair of primers, resulting in an increase in sensitivity and specificity. The product of the first amplification reaction is used as the template for the second, which is primed by oligonucleotides that are placed internal to the first primer pair. The use of two pairs of oligonucleotides allows for a higher number of cycles to be performed, thereby increasing the sensitivity of the PCR. Feng ZS et al. previously developed a novel locked nucleic acid (LNA)-based one-step single-tube nested (OSN)-qRT-PCR strategy to detect viral and bacterial pathogens with higher sensitivity and specificity than qRT-PCR and without the need of lid opening^[20]^. To improve the diagnostic accuracy of nucleic acid detection of SARS-CoV-2 in low viral load samples, they developed and evaluated the sensitivity and accuracy of OSN-qRT-PCR in detecting SARS-CoV-2^[21]^. However, the analytical performance of OSN-qRT-PCR in the published study is insufficient, lacking information such as specificity, reportable range, and the limit of detection (LoD). In addition, no studies have been conducted comparing the clinical application value of OSN-qRT-PCR and ddPCR for detecting SARS-CoV-2.

Here we provide a comparison of OSN-qRT-PCR and ddPCR with qRT-PCR using a dilution series of SARS-CoV-2 pseudoviral RNA and clinical samples. The detectable range and sensitivity of each assay were determined and clinical samples (n = 34) were used to validate clinical sensitivity and specificity. Compared with qRT-PCR and ddPCR, OSN-qRT-PCR showed higher sensitivity and greater practicality, making it better suited for detecting SARS-CoV-2 in low viral load samples.

## MATERIALS AND METHODS

### Ethics statement

The Ethics Committee of The Second People’s Hospital of Fuyang approved this study. Existing samples collected during standard diagnostic tests were tested and analyzed by qRT-PCR, OSN-qRT-PCR, and ddPCR. No extra burden was posed to patients.

### Specimen collection

We retrospectively identified 24 hospitalized patients clinically diagnosed with COVID-19 between January 30, 2020, and February 17, 2020, in The Second People’s Hospital of Fuyang. Throat (n = 18) and anal (n = 4) swabs, sputum (n = 10), and blood (n = 2) samples were collected from the enrolled patients. All aspects of the study were performed according to national ethics regulations and approved by the Institutional Review Boards of China Center for Disease Control and Prevention (CDC). Written consent was obtained from patients or children’s parents.

### SARS-CoV-2 pseudovirus preparation

The SARS-CoV-2 reference sequence was synthesized and cloned into a lentiviral vector and pseudovirus was prepared in 293T cells. The obtained pseudovirus contained RNA sequences of the *ORF1ab* and *N* genes in the lentiviral genome. The SARS-CoV-2 pseudovirus used in qRT-PCR and OSN-qRT-PCR was synthesized and processed by BDS company (DA’an, Guangzhou, China) at a RNA concentration of 2.0 x 10^4^ copies/ml. The SARS-CoV-2 pseudovirus used in ddPCR was synthesized and processed by BioPerfectus Technologies Co. (Taizhou, China) at a RNA concentration of 1.5 x 10^5^ copies/ml. The SARS-CoV-2 pseudoviral RNA was diluted with pseudovirus diluent (dilution ratio and method is shown in table 1), and SARS-CoV-2 pseudoviral RNA of the diluted samples S1, S2, S3, S4, S5, S6, S7, and S8 were extracted by using membrane adsorption kits (Di’an, Hangzhou, China).

**TABLE 1.** Dilution ratio and the concentration (copies/ml) of SARS-CoV-2 pseudovirus RNA standards used in each assay.

| qRT-PCR and OSN-qRT-PCR | ddPCR |
| --- | --- |

|  | Dilution ratio | Concentration<br>(copies/ml) | Dilution ratio | Concentration<br>(copies/ml) |
| --- | --- | --- | --- | --- |
| S0 | 0 | 20000 | 0 | 150000 |
| S1 | 5 | 4000 | 5 | 30000 |
| S2 | 10 | 2000 | 20 | 7500 |
| S3 | 20 | 1000 | 100 | 1500 |
| S4 | 40 | 500 | 300 | 500 |
| S5 | 100 | 200 | 600 | 250 |
| S6 | 200 | 100 | 1000 | 150 |
| S7 | 400 | 50 | / | / |
| S8 | 1000 | 20 | / | / |

### RNA extraction

Total RNA from throat and anal swabs, sputum, and blood samples from each patient was extracted from supernatants using Reagent of Nucleic Acid Extraction or Purification (Di’an, Hangzhou, China) following the manufacturer’s instructions. SARS-CoV-2 nucleic acid detection was mainly targeted at the two-segment conserved gene sequence of its genome, located at *ORF1ab* and *N*.

### ddPCR workflow

Reaction components of the ddPCR assay kit (BioPerfectus) included 5 μl of Supermix, 2 μl of reverse transcriptase, 1 μl of 300 mM DTT, 5 µl of SARS-CoV-2 reaction solution, and 7 μl template. All procedures followed the manufacturer’s instructions for the QX200 Droplet Digital PCR System using Supermix for the probe (no dUTP) (Bio-Rad, Hercules, USA). 20 μl of each reaction mix was converted to droplets with the QX200 droplet generator. Droplet-partitioned samples were then transferred to a 96-well plate, sealed, and cycled in a C1000 Touch Thermal Cycler (Bio-Rad) under the following cycling protocol: 50°C for 60 min and 95°C for 10 min, followed by 40 cycles of 95°C for 30 s and 56°C for 1 min, then 98°C for 10 min and 4°C hold. FAM *(ORF1ab)* and HEX *(N)* channels were selected to detect SARS-CoV-2. The cycled plate was then transferred and FAM and HEX channels read using the QX200 reader. Each run contained positive and negative controls. Samples were only considered positive when both FAM and HEX channels had signals.

### OSN-qRT-PCR workflow

Reaction components of the OSN-qRT-PCR assay kit (Sansure, Changsha, China) included 20 μl of template, 26 μl of reaction buffer, and 4 μl of the enzyme mixture. After vortexing and centrifugation, the reaction tube was transferred to the LightCycler 480 II Real-Time PCR System (Roche, Basel, Switzerland). The OSN-qRT-PCR amplification reaction contained the following steps: 50°C for 30 min, 95°C for 1 min, 20 cycles at 95°C for 30 s, 70°C for 40 s, and 72°C for 40s, followed by 40 cycles at 95°C for 15 s, 60°C for 30 s, and 25°C for 10 s of instrument cooling. FAM *(ORF1ab)* and ROX (N) channels were selected to detect SARS-CoV-2, and the VIC channel was chosen to detect the reference gene (human *ABL1)*. Each run contained positive and negative controls. FAM, HEX, and VIC channels all showed typical S-shaped amplification curves. The result was considered valid when the cycle threshold (Ct) value of the reference gene was ≤37. The result was considered positive when the Ct values of both target genes were ≤35 and negative when they were both >35. If only one of the target genes had a Ct value ≤35 and the other was >35, it was interpreted as a single-gene positive.

### qRT-PCR workflow

The qRT-PCR kit (DaAn Gene; Guangzhou, China) included 17 μl of SARS-CoV-2 NC reaction solution A, 3 μl of NC reaction solution B, and 5 μl of template. After vortexing and centrifugation, the reaction tube was transferred to the LightCycler 480 II Real-Time PCR System (Roche). The qRT-PCR amplification reaction contained the following steps: 50°C for 15 min, 95°C for 15 min, 45 cycles at 94°C for 15 s, and 55°C for 45 s. FAM (N) and VIC *(ORF1ab)* channels were selected to detect SARS-CoV-2, and the CY5 channel was chosen to detect the reference gene (human *ABL1)*. The result was considered valid when the Ct value of the reference gene was ≤37. The result was considered positive when the Ct values of both target genes *(ORF1ab* and *N)* were ≤37 and were considered negative when they were both >40. If only one of the target genes had a Ct value fall in the gray zone (37-40), it was retested. If the repeated result was positive for only one of two targets genes, it was interpreted as positive.

### Dynamic range and LoD of OSN-qRT-PCR, ddPCR, and qRT-PCR

To evaluate the dynamic range and consistency of OSN-qRT-PCR, ddPCR, and qRT-PCR, we first ran a serial dilution of the linear RNA standard for each assay. To determine the LoD, the lower concentration RNA standards (including S3-S8) were analyzed 14 times. The LoD was calculated by Probit regression analysis with a 95% repeatable probability.

### Data statistical analysis

Analysis of the ddPCR data was performed with Quanta Soft Analysis Software v1.7.4 to calculate the concentration of the target. Plots of linear regression were conducted with GraphPad Prism 7.0, and Probit analysis for LoD was conducted with MedCalc software v19.2.1. Bland-Altman analysis of qRT-PCR, OSN-qRT-PCR, and ddPCR results for patient samples was evaluated by SPSS 23.0 statistical software.

## RESULTS

### Comparison of the reportable range of each assay

To compare the reportable range of qRT-PCR, OSN-qRT-PCR, and ddPCR, the SARS-CoV-2 pseudoviral RNA standard was serially diluted from 2 × 10^4^ to 20 copies/ml for qRT-PCR and OSN-qRT-PCR, and from 1 × 10^5^ to 150 copies/ml for ddPCR. As shown in figure 1, the detectable range of qRT-PCR was 500 to 2 × 10^4^ copies/ml for *ORF1ab* and N, with R^2^ = 0.9985 and 0.9967, respectively (Fig. 1a,b). The detectable range of OSN-qRT-PCR was 100 to 2 × 10^4^ copies/ml for *ORF1ab* and 50 to 2 × 10^4^ copies/ml for N, with R^2^ = 0.9874 and 0.9936, respectively (Fig. 1c,d). Likewise, the detectable range of ddPCR was 250 to 1.5 × 10^5^ copies/ml for *ORF1ab* and N, with R^2^ = 0.9983 and 0.9984, respectively (Fig. 1e,f). These results show that the minimum detection range of OSN-qRT-PCR is significantly lower than those of qRT-PCR and ddPCR. Moreover, both OSN-qRT-PCR and ddPCR results displayed good linearity, suggesting that both assays can reliably detect SARS-CoV-2.

**Fig 1.**
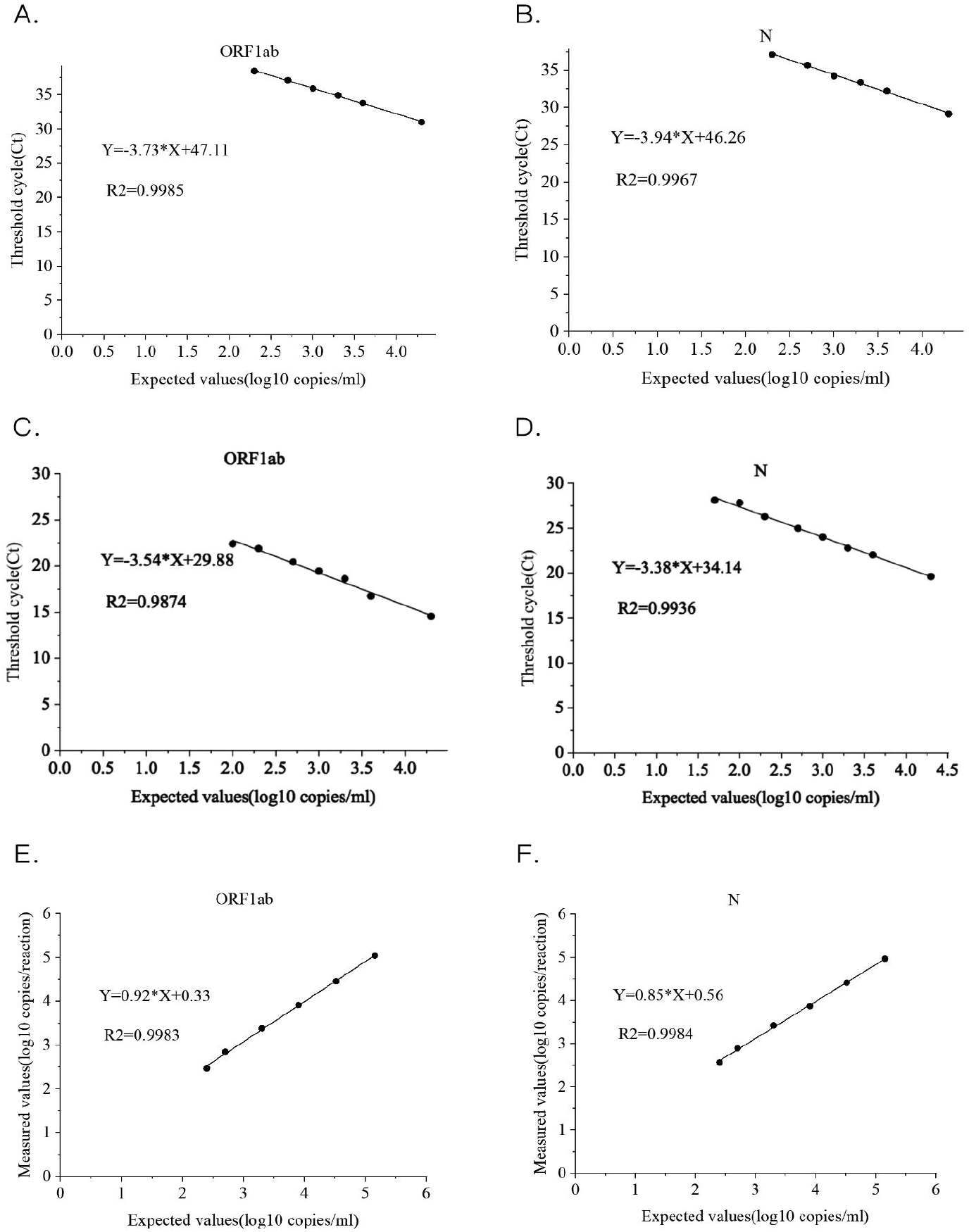
Linear relationship of qRT-PCR, OSN-qRT-PCR and ddPCR for quantifying SARS-CoV-2 pseudoviral RNA standards for *ORF1ab* and *N* gene. A. B: Expected values (converted to log^10^) were plotted on the X axis versus measured values of qRT-PCR (converted to log^10^) on the Y axis targeting (A) *ORF1ab* and (B) *N* C. D: Expected values(converted to log^10^) were plotted on the X axis versus measured Ct values of OSN-qRT-PCR on the Y axis targeting (C) *ORF1ab* and (D) N. E. F: Expected values(converted to log^10^) were plotted on the X axis versus measured values (converted to log^10^) of ddPCR on the Y axis using Graph Pad Prism targeting (C) *ORF1ab* and (D) N.

In addition, we compared the correlation between OSN-qRT-PCR and ddPCR with qRT-PCR and, as shown in figure 2, found that the Pearson correlation coefficients between qRT-PCR and OSN-qRT-PCR were 0.887 for *ORF1ab* and 0.742 for *N* (Fig. 2a,b), and -0.924 for *ORF1ab* and -0.844 for *N* between qRT-PCR and ddPCR (Fig. 2c,d). The good correlation between qRT-PCR with the other two assays further confirms that OSN-qRT-PCR and ddPCR are suitable methods for detecting SARS-CoV-2.

**Fig 2.**
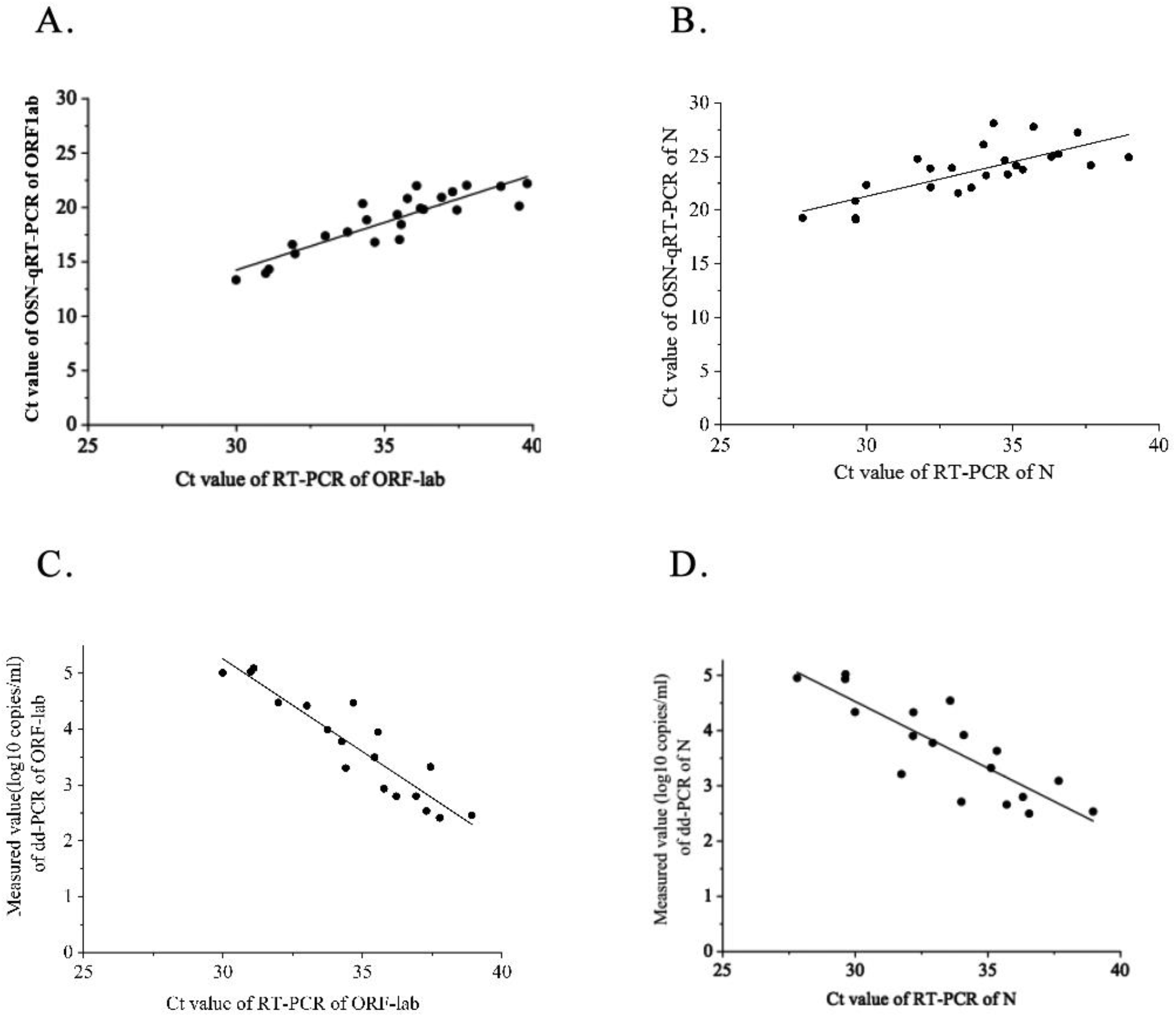
The correlation analysis between OSN-qRT-PCR and ddPCR with qRT-PCR for quantifying SARS-CoV-2 pseudoviral RNA standards for *ORF1ab* and *N* gene. A. B: the CT values of qRT-PCR of *ORF1ab* (A) and *Ngene* (B) were plotted on the X axis versus the CT values of OSN-qRT-PCR of *ORF1ab* and *N* gene on the Y axis, the Person Correlation Coefficient between qRT-PCR and OSN-qRT-PCR for *ORF1ab* and *N*gene were 0.887 and 0.742. C. D: the CT values of qRT-PCR of *ORF1ab* (C) and *Ngene* (D) were plotted on the X axis versus the measured values(converted to log^10^) of ddPCR of *ORF1ab* and *N gene* on the Y axis, the Person Correlation Coefficient between qRT-PCR and ddPCR for *ORF1ab* and *N* gene were -0.924 and -0.844.

### Comparison of the LoD for each assay

A variety of procedures are available for establishing the LoD for laboratory assays. The LoD is generally determined in one of two ways: either (i) statistically, by calculating the point at which a signal can be distinguished from background, or (ii) empirically, by testing serial dilutions of samples with a known concentration of the target substance in the analytical range of the expected detection limit. For medical applications of molecular assays, it is generally more meaningful to use the empirical method to estimate the detection limit.

The LoD was calculated by Probit regression analysis with a 95% repeatable probability, which is a commonly used method when empirically determining the limit of analyte that can be reliably detected. A series of linear SARS-CoV-2 pseudoviral RNA concentrations (including S3-S8) were prepared by diluting a high-concentration standard, with each concentration tested with 14 replicates. As shown in figure 3 and table 2, the LoD of qRT-PCR was 520.1 (95% confidence interval (CI): 363.23–1145.69) and 528.1 (95% CI: 347.7–1248.7) copies/ml for *ORF1ab* and N, respectively (Fig. 3a,b). The qRT-PCR kit claims a detection sensitivity of 500 copies/ml and was officially approved by the National Medical Products Administration (NMPA) and used for the detection of COVID-19 nationwide. Our LoD result was consistent with this claimed detection limit.

In contrast, the LoD of OSN-qRT-PCR was 194.74 (95% CI: 139.7–430.9) and 189.1 (95% CI: 130.9–433.9) copies/ml for *ORF1ab* and N, respectively (Fig. 3c,d), while the LoD of ddPCR was 401.8 (95% CI: 284.8–938.3) and 336.8 (95% CI: 244.6–792.5) copies/ml for *ORF1ab* and N, respectively (Fig. 3e,f). Taken together, these results show that the sensitivity of OSN-qRT-PCR is higher than both ddPCR and qRT-PCR, with ddPCR being more sensitive than qRT-PCR.

**Fig 3.**
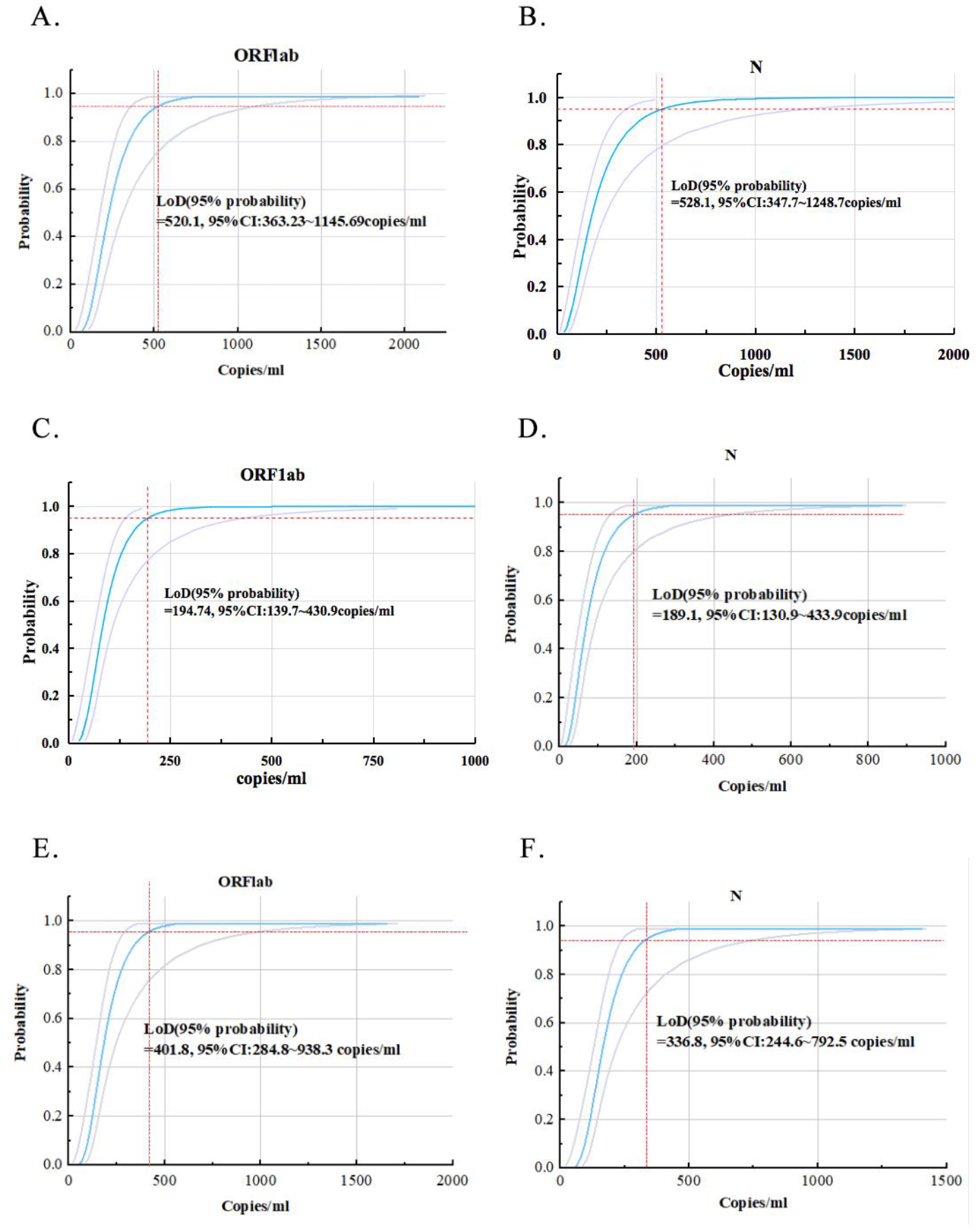
Probit analysis sigmoid curve reporting the LoD of each assay. Replicate reactions of *ORF1ab* (A) and *N* gene (B) of qRT-PCR, *ORF1ab* (C) and *N* gene (D) of OSN-qRT-PCR and *ORF1ab* (C) and *N* gene (D) of ddPCR were done at concentrations around the detection end point determined in preliminary dilution experiments. The X axis shows expected concentration (copies/ml). The Y axis shows fraction of positive results in all parallel reactions performed. The inner line is a probit curve (dose-response rule). The outer lines are 95% confidence interval (95% CI). Data are representative of three independent experiments with 14 replicates for each concentration.

**TABLE 2.**
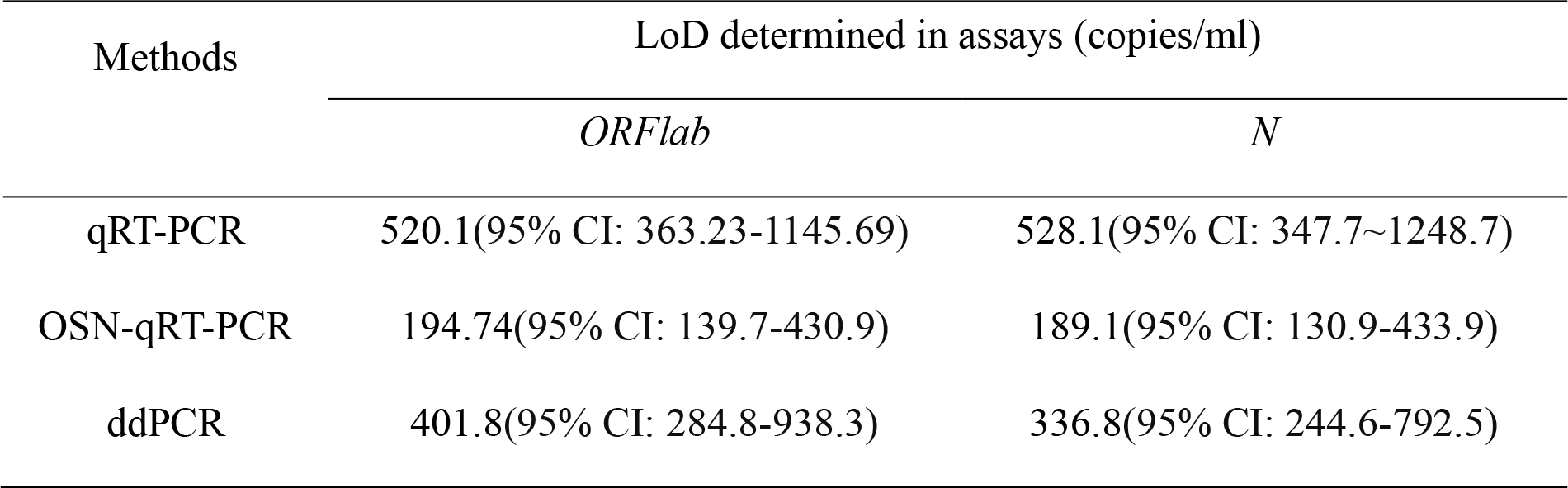
Estimated limit of detection for SARS-COV-2 in copies/ml for each assay.

| Methods | LoD determined in assays (copies/ml) |  |
| --- | --- | --- |
|  | *ORFlab* | *N* |
| qRT-PCR | 520.1(95% CI: 363.23-1145.69) | 528.1(95% CI: 347.7~1248.7) |
| OSN-qRT-PCR | 194.74(95% CI: 139.7-430.9) | 189.1(95% CI: 130.9-433.9) |
| ddPCR | 401.8(95% CI: 284.8-938.3) | 336.8(95% CI: 244.6-792.5) |

### Comparison of repeatability and conformity using intra-assay and inter-laboratory quality assessment panels

A series of linear SARS-CoV-2 pseudoviral RNA concentrations (including S0-S6) were tested in triplicates within the same run. As shown in table 3, the coefficient of variation (CV) values of intra-assays ranged from 1.60%-5.92% for qRT-PCR, 1.47%-4.99% for OSN-qRT-PCR, and 2.04%-11.18% for ddPCR. Overall, these results show that qRT-PCR and OSN-qRT-PCR assays demonstrate good repeatability. The cause of the unstable ddPCR results may be a consequence of differences in the number of microspheres due to poor machine operation by the experimenter.

**TABLE 3.** Comparison of the repeatability (CV%) of the each assay.

| Method | Conc.(copies/ml) | *ORFlab* |  |  | *N* |  |  |
| --- | --- | --- | --- | --- | --- | --- | --- |
|  |  | SD | MEAN | CV (%) | SD | MEAN | CV (%) |
| qRT-PCR | 20000 | 0.61 | 30.69 | 1.99% | 1.05 | 29.02 | 3.61% |
|  | 4000 | 1.36 | 33.22 | 4.09% | 1.81 | 31.92 | 5.67% |
|  | 2000 | 0.93 | 34.52 | 2.70% | 0.96 | 33.06 | 2.91% |
|  | 1000 | 1.55 | 35.76 | 4.32% | 2.02 | 34.07 | 5.92% |
|  | 500 | 0.58 | 36.30 | 1.60% | 1.86 | 36.00 | 5.16% |
|  | 200 | 0.84 | 37.99 | 2.20% | 1.69 | 37.08 | 4.56% |
| OSN-qRT-PCR | 20000 | 0.49 | 13.87 | 3.57% | 0.09 | 19.21 | 0.47% |
|  | 4000 | 0.83 | 16.65 | 4.97% | 0.13 | 22.19 | 0.61% |
|  | 2000 | 1.35 | 18.85 | 7.15% | 0.38 | 23.68 | 1.62% |
|  | 1000 | 0.46 | 19.33 | 2.36% | 0.50 | 24.23 | 2.06% |
|  | 500 | 0.56 | 20.56 | 2.71% | 0.97 | 25.09 | 3.86% |
|  | 200 | 0.32 | 21.81 | 1.47% | 1.31 | 26.30 | 4.99% |
| ddPCR | 143000 | 5644.4 | 107327.3 | 5.26% | 2862.1 | 86757.2 | 3.30% |
|  | 33000 | 1999.9 | 28569.7 | 7.00% | 1902.0 | 22665.4 | 8.39% |
|  | 8000 | 458.7 | 9189.7 | 4.99% | 164.9 | 8094.9 | 2.04% |
|  | 2000 | 75.8 | 2085.0 | 3.63% | 340.8 | 4009.4 | 8.50% |
|  | 500 | 51.2 | 671.7 | 7.62% | 62.3 | 557.1 | 11.18% |
|  | 250 | 22.4 | 261.7 | 8.55% | 37.8 | 371.4 | 10.18% |

The inter-laboratory quality assessment panel provided by the National Center for Clinical Laboratories (NCCL) is used for evaluation of a laboratory’s ability to detect nucleic acids of SARS-CoV-2. This panel includes a total of 10 samples named as 202001-202010. As shown in table 4, the test results of each assay using this panel indicate that both OSN-qRT-PCR and ddPCR assays showed 100% specificity for detecting SARS-CoV-2 and were negative for other human coronaviruses.

**TABLE 4.** Comparison of test results and return results of inter-laboratory quality assessment panel.

| Sample ID | Test Results |  |  |  |  |  | Return Results |  |
| --- | --- | --- | --- | --- | --- | --- | --- | --- |
|  | qRT-PCR (Ct Value) |  | OSN-qRT-PCR (Ct Value) |  | ddPCR (copies/mL) |  | SARS-COV-2 | Pass Rate (%) |
|  | *ORF-1ab* | *N* | *ORF-1ab* | *N* | *ORF-1ab* | *N* |  |  |
| 202001 | — | — | — | — | — | — | negative | 99 |
| 202002 | 31.79 | 30.49 | 16.45 | 20.97 | 3074 | 2073 | positive | 98 |
| 202003 | 25.99 | 24.23 | 13.78 | 18.68 | 23080 | 5005 | positive | 99 |
| 202004 | 33.87 | 32.31 | 20.94 | 21.19 | 1144 | 2860 | positive | 98 |
| 202005 | 29.93 | 28.87 | 15.56 | 19.53 | 7293 | 2072 | positive | 99 |
| 202006 | — | — | — | — | — | — | negative | 99 |
| 202007 | 32.85 | 31.73 | 18.38 | 20.80 | 1144 | 1573 | positive | 98 |
| 202008 | — | — | — | — | — | — | negative | 99 |
| 202009 | 34.85 | 33.91 | 18.92 | 22.43 | 1312 | 643 | positive | 97 |
| 202010 | 27.79 | 26.40 | 12.22 | 15.89 | 33962 | 11072 | positive | 99 |

### Comparison of each assay for SARS-CoV-2 RNA detection using patient samples in the acute phase of infection

A total of 34 samples from 24 COVID-19-confirmed patients diagnosed in the acute phase of infection were analyzed by each assay. The positive threshold for SARS-CoV-2 detection was defined as values equal to or greater than the LoD of *ORF1ab* and *N* primers/probe sets. The results of qRT-PCR, OSN-qRT-PCR, and ddPCR for each sample are shown in tables 5 and 6. Among the 34 samples, 14 samples were initially qRT-PCR negative (positive rate = 58.82%), while 28 tested positive by OSN-qRT-PCR (positive rate = 82.35%). In addition, 23 tested positive by ddPCR (positive rate = 67.65%), indicating a higher sensitivity than qRT-PCR but lower than OSN-qRT-PCR. These results were further analyzed by the Bland-Altman method, which reveals the agreement between two independent methods. As shown in figure 4, OSN-qRT-PCR and ddPCR results were in good agreement with qRT-PCR. Therefore, both OSN-qRT-PCR and ddPCR assays proved capable of detecting SARS-CoV-2 in patient specimens.

**TABLE 5.** The results of qRT-PCR, OSN-qRT-PCR and ddPCR and further clinical information of 34 clinical samples from 24 acute phase COVID-19 patients.

| Sample ID. | Results of qRT-PCR<br>(Ct Value) |  | Results of OSN-qRT-PCR(Ct Value) |  | Results of ddPCR<br>(copies/ml) |  | Specimen type | Initial official reports by OSN-qRT-PCR | Judgment by N-PCR | Judgment by ddPCR |
| --- | --- | --- | --- | --- | --- | --- | --- | --- | --- | --- |
|  | *ORF1ab* | *N* | *ORF1ab* | *N* | *ORF1ab* | *N* |  |  |  |  |
| 1 | 27.20 | 26.94 | 15.00 | 16.68 | 129415 | 1287.00 | Throat swab | P | P | P |
| 2 | 35.04 | 35.35 | 22.60 | 22.80 | 428.90 | 572.00 | Throat swab | P | P | P |
| 3 | — | — | 37.11 | 37.22 | — | — | Throat swab | N | N | N |
| 4 | 34.46 | 34.95 | 21.74 | 22.76 | 500.50 | 715.00 | Throat swab | P | P | P |
| 5 | 31.31 | 30.66 | 17.39 | 19.21 | 5148.00 | 10224.50 | Throat swab | P | P | P |
| 6 | 28.02 | 27.38 | 14.65 | 15.25 | 74360.0 | 364.45 | Throat swab | P | P | P |
| 7 | — | — | 38.21 | — | — | — | Throat swab | N | N | N |
| 8 | 32.61 | 32.70 | 18.17 | 19.96 | 2860.00 | 3503.50 | Throat swab | P | P | P |
| 9<sup>a</sup> | 40.32 | — | 23.50 | 24.64 | 228.80 | 429.00 | Throat swab | N | P | S |
| 10 | 35.50 | 35.10 | 23.40 | 23.96 | 715.00 | 715.00 | Throat swab | P | P | P |
| 11 | 35.08 | 33.09 | 23.60 | 22.14 | 715.00 | 2145.00 | Throat swab | P | P | P |
| 12 | 26.58 | 26.76 | 12.52 | 13.95 | 215930 | 4336.0 | Throat swab | P | P | P |
| 13<sup>a</sup> | 40.83 | — | 28.75 | 26.19 | 157.30 | 228.80 | Throat swab | N | P | N |
| 14 | 34.00 | 33.96 | 20.84 | 23.19 | 1930.50 | 1716.00 | Throat swab | P | P | P |
| 15 | 34.37 | 34.38 | 21.70 | 23.10 | 2216.50 | 1144.00 | Throat swab | P | P | P |
| 16 | — | — | — | 38.56 | — | — | Throat swab | N | N | N |
| 17 | 34.33 | 34.27 | 22.08 | 20.28 | 1215.50 | 2502.50 | Throat swab | P | P | P |
| 18<sup>a</sup> | — | 40.21 | 28.67 | 26.88 | 300.30 | 786.50 | Throat swab | N | P | S |
| 19 | 29.29 | 29.42 | 18.21 | 18.77 | 31460 | 42900 | sputum | P | P | P |
| 20 | — | — | 36.11 | — | 572.00 | 1787.50 | sputum | N | N | P |
| 21 | 29.89 | 29.25 | 18.30 | 19.36 | 13799 | 30530 | sputum | P | P | P |
| 22 | 30.45 | 29.82 | 21.82 | 23.99 | — | — | sputum | P | P | N |
| 23 | 26.79 | 26.94 | 15.78 | 17.50 | 23595.0 | 14657.50 | sputum | P | P | P |
| 24<sup>a</sup> | 40.81 | — | 35.98 | 36.21 | — | 1430.0 | sputum | N | N | N |
| 25<sup>a</sup> | 40.21 | — | 27.88 | 25.63 | 643.50 | 1215.50 | sputum | N | P | P |
| 26 | 31.10 | 30.32 | 22.56 | 21.94 | 9509.50 | 3789.50 | sputum | P | P | P |
| 27 | 27.85 | 27.45 | 20.67 | 20.34 | 84370.0 | 170170.0 | sputum | P | P | P |
| 28 | 29.89 | 28.22 | 22.10 | 21.31 | 643.50 | — | sputum | P | P | N |
| 29<sup>a</sup> | — | 40.22 | 16.62 | 17.08 | 715.0 | 715.0 | blood | N | P | P |
| 30 | — | — | — | 37.90 | — | — | blood | N | N | N |
| 31 | 35.08 | 34.73 | 21.57 | 22.83 | 500.50 | 715.00 | Anal swab | P | P | P |
| 32<sup>a</sup> | — | 40.55 | 25.20 | 24.68 | 214.50 | — | Anal swab | N | P | N |
| 33<sup>a</sup> | — | 40.06 | 24.00 | 25.68 | — | 250.25 | Anal swab | N | P | N |
| 34 | — | — | 26.66 | 27.22 | 307.45 | — | Anal swab | N | P | N |

a: one or two of the target genes had a Ct value happen to fall in the gray zone (Ct value: 37-40), and samples were retested by qRT-PCR. At last, these samples were considered as negative with Ct value>40 (both targets genes) by qRT-PCR.
P, positive; N, Negative; S, suspect; GGO, ground glass opacities image.

**TABLE 6.** Reports summary of qRT-PCR, N-PCR and ddPCR for 34 clinical samples.

|  | qRT-PCR | OSN-qRT-PCR | ddPCR |
| --- | --- | --- | --- |
|  | 20 P | 28 P | 21P |
| 34 |  |  |  |
| Samples | 14 N | 6 N | 2 S |

P, positive; N, Negative; S, suspect;

**Fig 4.**
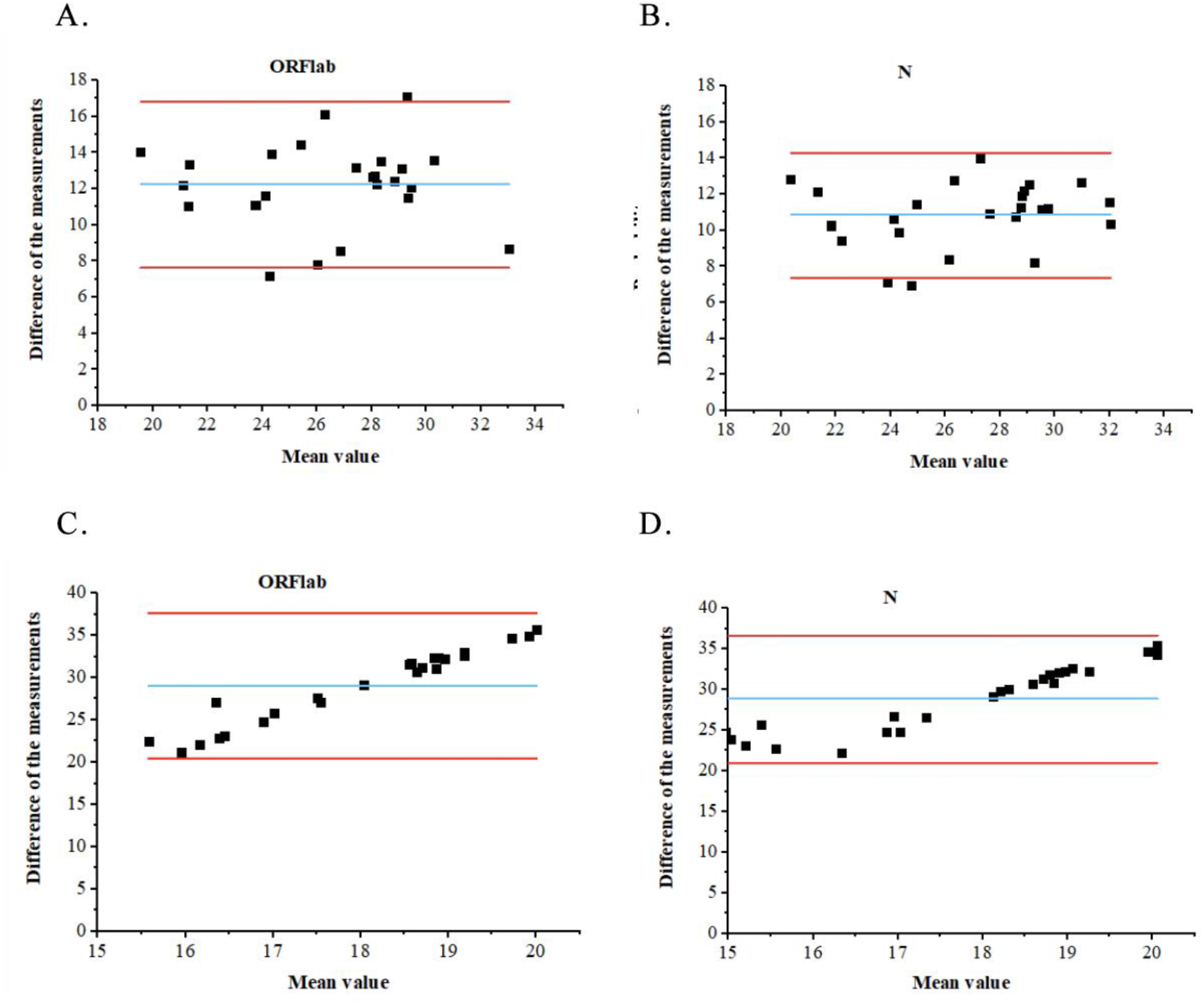
Bland-Altman plots of SARS-CoV-2 quantification by using three methods in patient specimens. A. B: Bland-Altman plots comparing qRT-PCR and OSN-qRT-PCR assays for patient specimens. C. D: Bland-Altman plots comparing qRT-PCR and ddPCR assays for patient specimens. Notes: Blue lines indicate mean difference, Red lines indicate limits of agreement (LoA).

### Comparison of the positive rates of different specimen types from COVID-19 patients

A previous study revealed that SARS-CoV-2 exists in both the upper and lower respiratory tract^[22]^. We collected simultaneous sputum and throat swabs from a total of 10 COVID-19-confirmed patients diagnosed in the acute phase of infection and the three methods were used to detect SARS-CoV-2 nucleic acid. As shown in table 7, the positive rates in sputum samples for all three methods were significantly higher than those of throat swabs. In addition, we also collected blood specimens (n = 2) and anal swab specimens (n = 4) from COVID-19-confirmed patients to analyze the detection ability of the assays for other specimen types. As shown in table 4, for blood specimens (29# and 30#), qRT-PCR and ddPCR results were both negative, while the OSN-qRT-PCR result was positive. For anal swab specimens (31#-34#), only one case was positive by ddPCR and qRT-PCR assay, while all four were positive by OSN-qRT-PCR. It is evident from these results that among the three methods, OSN-qRT-PCR has the greatest sensitivity for detecting SARS-CoV-2 from different specimen types.

**TABLE 7.** Comparison of the positive rate of SARS-CoV-2 nucleic results from different specimen types from same patient at same time.

|  | qRT-PCR | OSN-qRT-PCR | ddPCR |
| --- | --- | --- | --- |
| Throat swab | 50% | 70% | 50% |
| (n=10) | (5/10) | (7/10) | (5/10) |
| Sputum | 70% | 80% | 70% |
| (n=10) | (7/10) | (8/10) | (7/10) |

## DISCUSSION

The clinical detection sensitivity of qRT-PCR is affected by various factors, such as the nucleic acid extraction method, the one-step qRT-PCR reagent used, and the primer/probe sets^[23]^. It has been reported that seven commercial qRT-PCR detection kits revealed significant differences in the detection ability for weakly positive samples^[24]^. ddPCR has exhibited higher sensitivity and precision than classical qRT-PCR^[25,26]^. Recent studies have confirmed that both ddPCR and OSN-qRT-PCR are strongly recommended in clinical practice for the diagnosis of COVID-19 and for follow-up of positive patients until complete remission^[21,27]^. However, ddPCR is limited to special equipment, which hinders its clinical application. Compared with ddPCR, the advantage of OSN-qRT-PCR is greater practicality because of easier adaptation for laboratories already equipped with traditional real-time PCR machines.

Here, for the first time, we provide a head-to-head comparison of OSN-qRT-PCR and ddPCR with qRT-PCR for the detection of SARS-CoV-2 using a pseudoviral RNA standard, inter-laboratory quality assessment panel, and clinical samples of different types. The detectable range, consistency, specificity, and LoD of each method were comparably analyzed. Our results demonstrate that OSN-qRT-PCR and ddPCR are reliable for quantitatively detecting SARS-CoV-2. In addition, Bland-Altman analysis showed that ddPCR and OSN-qRT-PCR had good correlation with qRT-PCR in testing clinical specimens. In particular, the detection performance of both OSN-qRT-PCR and ddPCR assays were better than qRT-PCR, and the OSN-qRT-PCR assay had the lowest LoD, suggesting that OSN-qRT-PCR and ddPCR assays are valuable additions for detecting SARS-COV-2 in samples with low viral loads. Although the sensitivity of OSN-qRT-PCR was reported to be 10-fold higher than qRT-PCR using plasmids^[21]^, our results revealed that the sensitivity of OSN-qRT-PCR was only 2-3-fold higher than qRT-PCR when using pseudoviral RNA.

A previous study revealed that SARS-CoV-2 exists in both the upper and lower respiratory tract and that the viral load in sputum is higher than that of throat swabs ^[28]^. Our findings also confirmed that although SARS-CoV-2 can colonize the upper respiratory tract, lower respiratory tract samples better reflect the viral replication level in infected patients. Although OSN-qRT-PCR and ddPCR both exhibited higher sensitivity than qRT-PCR, there were still false negative results (six missed by OSN-qRT-PCR, 11 missed by ddPCR) when analyzing clinical specimens. This may have been due to the quality of sample collection or viral loads falling below detection limits resulting from missing the optimum sample collection time. For specimen 20#, the results of OSN-qRT-PCR and qRT-PCR were negative, while ddPCR was positive. This discordant result may have been due to the specimen type (sputum) or the various influencing factors in the nucleic acid extraction process, leading to poor stability of test results.

This study had several limitations. First, the SARS-CoV-2 pseudoviral RNA concentration used in the study for exploring detectable ranges did not include high concentrations. Second, the clinical specimens were only from COVID-19-confirmed patients in the acute phase of infection; clinical specimens from patients in the recovery phase or suspected patients were not included. Finally, our study was limited by a small sample size and thus conclusions should be interpreted with caution and confirmed by further studies.

## CONCLUSION

We validated the implementation of OSN-qRT-PCR and ddPCR systems as new alternatives to qRT-PCR for the sensitive and accurate quantification of SARS-CoV-2, especially in samples with low viral loads. Considering its sensitivity and practicality, OSN-qRT-PCR is a highly valuable and feasible method that offers the potential to facilitate clinical diagnoses and decision-making for patients with COVID-19.

## Data Availability

Yes

## CONFLICTS OF INTEREST

The authors declare no conflicts of interests.

## FUNDING

This study was supported by the New Coronavirus Infection Emergency Science and Technology Project, Clinical Research Hospital of Chinese Academy of Sciences[grant number YD9110002010], the Fundamental Research Funds for the Central University [grant number WK9110000025], the Natural Science Foundation of Anhui Province (Grant Number: 2008085MH288), the China Mega-Projects for Infectious Disease (2017ZX10104001, 2018ZX10711001, and 2018ZX10713-002).

## Notes

### Competing Interest Statement

The authors have declared no competing interest.

### Clinical Trial

JCM02192-20

### Author Declarations

MEDRXIV/2020/182832

## REFERENCE

[1] World Health Organization (WHO), COVID-19. https://www.who.int/emergencies/diseases/novel-coronavirus-2019, 2020 (accessed 8 April 2020).

[2] V.M. Corman, O. Landt, M. Kaiser, R. Molenkamp, A. Meijer, D.K.W. Chu, T. Bleicker, S. Brünink, J. Schneider, M.L. Schmidt, D.G.J.C. Mulders, B.L. Haagmans, B. van der Veer, S. van den Brink, L. Wijsman, G. Goderski, J.L. Romette, J. Ellis, M. Zambon, M. Peiris, H. Goossens, C. Reusken, M.P.G. Koopmans, C. Drosten, Detection of 2019 novel coronavirus (2019-nCoV) by real-time RT-PCR, Euro. Surveill. 25 (2020).

[3] K. Shirato, N. Nao, H. Katano, I. Takayama, S. Saito, F. Kato, H. Katoh, M. Sakata, Y. Nakatsu, Y. Mori, T. Kageyama, S. Matsuyama, M. Takeda, Development of Genetic Diagnostic Methods for Novel Coronavirus 2019 (nCoV-2019) in Japan, Jpn. J. Infect. Dis. (2020).

[4] R. Konrad, U. Eberle, A. Dangel, B. Treis, A. Berger, K. Bengs, V. Fingerle, B. Liebl, N. Ackermann, A. Sing, Rapid establishment of laboratory diagnostics for the novel coronavirus SARS-CoV-2 in Bavaria, Germany, February 2020, Euro. Surveill. 25 (2020).

[5] D.K.W. Chu, Y Pan, S.M.S. Cheng, K.P.Y. Hui, P. Krishnan, Y Liu, D.Y.M. Ng, C.K.C. Wan, P. Yang, Q. Wang, M. Peiris, L.L.M. Poon, Molecular Diagnosis of a Novel Coronavirus (2019-nCoV) Causing an Outbreak of Pneumonia, Clin. Chem. 66 (2020) 549-555.

[6] Winichakoon, P.; Chaiwarith, R.; Liwsrisakun, C.; Salee, P.; Goonna, A.; Limsukon, A.; Kaewpoowat, Q., Negative Nasopharyngeal and Oropharyngeal Swab Does Not Rule Out

[7] Wu J, Liu J, Zhao X, et al. Clinical characteristics of imported cases of COVID-19 in Jiangsu Province: a multicenter descriptive study. Clin Infect Dis 2020. doi:10.1093/cid/ciaa199.

[8] Vogelstein B, Kinzler KW. Digital PCR. Proc. Natl. Acad. Sci. U. S. A. 1999;96:9236-9241.

[9] Nyaruaba, R.; Mwaliko, C.; Kering, K.K.; Wei, H. Droplet digital PCR applications in the tuberculosis world. Tuberculosis 2019, 117, 85-92.

[10] Green M R, Sambrook J. Nested Polymerase Chain Reaction (PCR)[J]. Cold Spring Harbor Protocols, 2019, 2019(2).

[11] Pohl G, Shih I-M. Principle and applications of digital PCR. Expert Rev. Mol. Diagn. 2004;4:41-47.

[12] Sanders R, Mason DJ, Foy CA, et al. Evaluation of Digital PCR for Absolute RNA Quantification. PLoS One. 2013;8:e75296.

[13] White RA, Blainey PC, Fan HC, et al. Digital PCR provides sensitive and absolute calibration for high throughput sequencing. BMC Genomics. 2009;10:110-116.

[14] Hindson CM, Chevillet JR, Briggs HA, et al. Absolute quantification by droplet digital PCR versus analog real-time PCR. Nat. Methods. 2013;10:1003-1005.

[15] Brunetto GS, Massoud R, Leibovitch EC, et al. Digital droplet PCR (ddPCR) for the precise quantification of human T-lymphotropic virus 1 proviral loads in peripheral blood and cerebrospinal fluid of HAM/TSP patients and identification of viral mutations. J. Neurovirol. 2014;20:341-351.

[16] Caviglia GP, Abate ML, Tandoi F, et al. Quantitation of HBV cccDNA in anti-HBc-positive liver donors by droplet digital PCR: A new tool to detect occult infection. J. Hepatol. [Internet]. 2018;69:301-307. Available from: https://doi.org/10.1016/jjhep.2018.03.021.

[17] Postel M, Roosen A, Laurent-Puig P, et al. Droplet-based digital PCR and next generation sequencing for monitoring circulating tumor DNA: a cancer diagnostic perspective. Expert Rev. Mol. Diagn. 2018;18:7-17.

[18] Zhao, Jiankang & Li, Haibo & Li, Hui & Wu, Qiaoling & Wu, Ke & Xiong, Zhujia & Yu, Zhongguang & Zhu, Yue & Fan, Yanyan & Li, Binbin & Ye, Yufei & Lu, Binghuai & Cao, Bin. (2020). Viral load in upper respiratory tract of COVID-19 patients detected by digital PCR. 10.21203/rs.3.rs-29834/v1.

[19] Lu R, Wang J, Li M, Wang Y, Dong J, Cai W. SARS-CoV-2 detection using digital PCR for COVID-19 diagnosis, treatment monitoring and criteria for discharge. medRxiv preprint. 2020;Available from: https://doi.org/10.1101/2020.03.24.20042689.

[20] Feng Z S, Zhao L, Wang J, et al. A multiplex one-tube nested real time RT-PCR assay for simultaneous detection of respiratory syncytial virus, human rhinovirus and human metapneumovirus[J]. Virology Journal, 2018, 15(1).

[21] Ji W, Kun C, Ruiqing Z et al. A novel one-step single-tube nested quantitative Real-Time PCR assay for highly sensitive detection of SARS-CoV-2[J]. Anal Chem, DOI:10.1021/acs.analchem.0c01884

[22] Chan JF, Yuan S, Kok KH, et al. A familial cluster of pneumonia associated with the 2019 novel coronavirus indicating person-to-person transmission: a study of a family cluster. Lancet 2020;395:514-23.

[23] Xie, X.; Zhong, Z.; Zhao, W.; Zheng, C.; Wang, F.; Liu, J., Chest CT for Typical 2019-nCoV Pneumonia: Relationship to Negative RT-PCR Testing. Radiology 2020, 200343.

[24] Kasteren P B V, Veer B V D, Brink S V D, et al. Comparison of seven commercial RT-PCR diagnostic kits for COVID-19[J]. Journal of Clinical Virology, 2020.

[25] Strain MC, Lada SM, Luong T, Rought SE, Gianella S, Terry VH, et al. Highly Precise Measurement of HIV DNA by Droplet Digital PCR. Wu Y, éditeur. PLoS ONE. 2013;8(4):e55943.

[26] Huang J-T, Liu Y-J, Wang J, Xu Z-G, Yang Y, Shen F, et al. Next Generation Digital PCR Measurement of Hepatitis B Virus Copy Number in Formalin-Fixed Paraffin-Embedded Hepatocellular Carcinoma Tissue. Clinical Chemistry. 1 janv 2015;61(1):290–6.

[27] Falzone L, Musso N, Gattuso G, et al. Sensitivity assessment of droplet digital PCR for SARS-CoV-2 detection[J]. International Journal of Molecular Medicine, 2020.

[28] Pan Y, Zhang D, Yang P, Poon LLM, Wang Q. Viral load of SARS-CoV-2 in clinical samples. Lancet Infect Dis 2020; 20:411-2.

